# Pooled Estimate of Risky Sexual Behavior among college and university students in sub-Saharan Africa: A Meta-Analysis

**DOI:** 10.1101/2022.05.28.22275722

**Authors:** A. Lungu, C. Chella, M. Zambwe, P.J. Chipimo

**Author notes:** **<u>CORRESPONDING AUTHOR DETAILS</u> Name:** Mowa Zambwe, **Address:** Workers Compensation Fund Control Board, Lusaka, Zambia, **Phone Number:** **, **Email ID:**. **Guarantor of Submission:** *The corresponding author is the guarantor of submission.

## Abstract

**Objective:** To determine the pooled estimate of risky sexual behaviour among university students in sub Saharan Africa.

**Methods:** A meta analytic study conducted to identify predictors of risky sexual behavior among university students. Databases from PubMed, African Journals Online, Science Direct, Google Scholar were used to identify appropriate studies. The combined effect estimates for each outcome were computed in Meta XL using random effects.

**Results:** The estimated pooled prevalence of sexual activity among university students was 51.0% (95% CI: 43.0% - 59.0%). Pooled prevalence for multiple sexual partners was 36.0% (95% CI: 30.0% - 42.0%), inconsistent condom use, 53.0% (95% CI: 46% - 61.0%) and for at least one risky sexual behavior, 65.0% (48.0% - 81.0%). Males were 3.36 times [OR: 3.05; 95% CI: 2.59 - 4.37] more likely to have multiple sexual partners than females. This review also indicated that males were 2.99 times [OR: 2.99; 95% CI: 1.40 - 6.40] more likely to engage in at least one risky sexual behavior than females.

**Conclusion:** Inconsistent condom use and multiple sexual partners were the most rampant risky sexual behaviors in Universities in Sub-Saharan Africa. Sustained risk communication on Sexual and Reproductive Health and youth friendly programs are highly recommended.

## BACKGROUND

Risky Sexual Behavior (RSB) is an act that increases one’s risk of contracting sexually transmitted infections and experiencing unintended pregnancy [1]. It includes multiple sexual partners, a history of unprotected sex/failure to use condoms or intermittent use, exchanging money for sex, performing sexual intercourse while under the influence of alcohol [1,2]. Adolescents constitute the largest percentage of people in developing countries, especially in sub-Saharan Africa. Promotion of safe sex and encouragement of contraceptive use would contribute immensely to the reduction in sex-related morbidity and mortality caused by teenage pregnancy, abortion, HIV/AIDS and, at the same time, reduce the population explosion)[3].

Young people aged 15–19 years have been reported to engage in risky sexual behaviours such as early sexual debut, multiple sexual partnerships and inconsistent condom use [4,5,6]. Nearly 33% of new HIV infections occur in young people between 15–24 years [7,8]. Most university students are in the youth age category and are categorized under the most at-risk population group due to their inclination to be engaged in risky sexual behavior, which leads to acquiring STIs [9,10,11]. Studies have indicated that risky sexual behaviors are fueling the spread of HIV and AIDS and unwanted pregnancy among young people in Sub-Saharan Africa [16,17].

Studies reviewed in Sub-Saharan Africa reveal that most university and college students are sexually activity with then beginning sexual intercourse during adolescence. A study conducted in Cameroon among 411 university students found that 80.8 % of students were sexually active with a mean age at sexual debut at 18.1 years (SD = 3.1). The frequency of premarital sex was 92.8 % [18]. Similarly, a large cross-sectional study among college students in Gauteng and North West Provinces of South Africa found that of the 3,674 students who participated in the survey, 2,947 (80.7%) reported being sexually active, and 2,408 (73.6%) had sex in the last three months [19]. Compared to the preceding two studies, a cross-sectional study conducted among 427 college students at the University of Zambia reports a much lower prevalence of sexual activity. The study found that 205 (48%) students reported having sex [20]. A similar study reveals that the proportion of early sexual initiation among college students was 17.9% [95% CI: 14.4%-24.4%] [21].

Similarly, a cross-sectional study in Ibadan of Nigeria reported that the majority (65.3%) were sexually active in the last 12 months [22]. Additionally, a study conducted in Ethiopia revealed that close to 3 in 10 (28.34%) of the total study participants reported having had sexual intercourse at least once. More proportion of male students ever had sex compared with females. One-fifth of these students had their first sexual experience after they joined university [23].

In a descriptive cross-sectional study conducted among 300 undergraduates in Port Harcourt, Nigeria, reports that more than half (52.0%) of the respondents had either boy/girlfriend, and a total of 144 (52.0%) had ever had sexual intercourse). Similarly, over 40% (48.6%) of respondents in Nigerian universities were currently sexually active, with the mean age at sexual debut at 17.0± 4.5years [24].

Another similar study conducted in South Africa among 1060 university students in Mahikeng discovered that the average age of first sexual intercourse for females (18 years) was higher than the average for males (18 years) (16 years)[25].

In a South African based study conducted among 576 students, 218 (37.8%) (83 women, 135 men) reported vaginal intercourse in the past two months. Of these, 7% of women and 43% of men reported past-year concurrent partnerships, and 24% knew/suspected partner non-monogamy [26]. Similarly, a facility-based cross-sectional study design conducted among 797 regular undergraduate students of Mekelle University reported that overall, approximately 44% of the participants who had sex in the last 12 months practiced unsafe sex [27]. Significant reasons for unsafe sex were fun, lack of awareness, trusting sexual partners, and no access to a condom [27].

The study of risky sexual behaviour among 1,286 undergraduate students at Haramaya University in Ethiopia from March to April 2010 observed a much lower prevalence of sexual activity when compared to the majority of the studies reviewed. Only 355 (28%; 95% CI 25.5-30.5) students reported having had sexual intercourse at least once. One fifth (22.8%) of these students had their sexual debut after they joined university. About six per cent of students with sexual experience reported having had intercourse with same-sex partners [28].

A descriptive cross-sectional survey that used multistage sampling to select 918 students also reports that with regards to sexual activity, males (54.1%) were more likely to be in a sexual relationship when compared to females (47.3%) [29].

We could not find meta-analysis, within the last 5 years, which aimed at consolidating the findings and provide estimates of risky sexual behaviors in the sub-Saharan region. Thus this study aimed to establish the pooled estimate of risky sexual behaviour among university and college students in sub-Saharan Africa.

## METHODS

A meta-analysis study involved primary quantitative articles published in the past ten years (2012 – 2021) in Sub-Saharan Africa.

PubMed, African Journals Online, Science Direct and Google Scholar were searched using the following terms and keywords:

“prevalence OR epidemiology OR magnitude OR incidence AND risky sexual behavior OR risky behavior AND college OR higher institution OR university AND students OR student OR learner OR learners AND” Low income” OR “Least developed country* OR “Low to middle income” OR “Under developed nation” OR “sub-Saharan Africa” OR “poor country*” OR “Third world country*” OR “Global south” OR “LMIC” OR MH “Developing countries” OR MH “Africa south of the Sahara”.” A similar approach was applied with Google Scholar, using the specific subject heading as advised for each database.

### Data extraction

The data extraction was done using a data extraction tool which included the title, author, year of survey and publication, study design, sample size, data collection procedure, study participants, study area, response rate, sampling method, and definition of risky sexual behavior.

### Publication bias and heterogeneity

The Joanna Briggs Institute Meta-Analysis of Statistics Assessment and Review Instrument (JBI-MAStARI) was used for critical appraisal. This tool includes a different appraisal checklist for each study design type. This instrument was used to assess articles before their inclusion in the final review by two independent reviewers. Any discrepancies between the reviewers were resolved via discussion and the involvement of a third reviewer. The final review included studies with a quality assessment score of at least 50% and a response rate of at least 80%.

### Statistical methods and analysis

The meta-analysis was conducted using Meta XL. Forest plots were used to present the combined estimate with 95% confidence intervals (CI).

## RESULTS

### Results of Literature Search

Our initial search yielded 2512 articles (Pubmed central = 2602; Other Sources = 130). Several articles at this stage were excluded from our studies because they were duplicates (n = 6) and did not meet our inclusion criteria (n = 2671), which left us with 55 articles as indicated through their titles. The screening process, which included scheming through the abstracts, further narrowed our potential studies for inclusion to 28 articles only. Several studies (n = 27) were excluded during this process, including those that utilized qualitative study designs, focused on youths and adolescents in informal settings, were from places other than Africa and studies whose full articles were not available. A total of 28 studies met our inclusion criteria for this particular study.

### Description of Studies Included

The review included 28 studies (18495 participants) carried out in Sub-Saharan Africa at selected colleges and Universities. Most of the studies included in the review were from Ethiopia (n = 18), followed by Nigeria (n = 3), South Africa (n = 3), Kenya (n = 1), Togo (n = 1), Zambia (n = 1) and Cameroon (n = 1). Most of these studies were carried out in universities (n = 20) compared to colleges (n = 8).

**Table 1.**
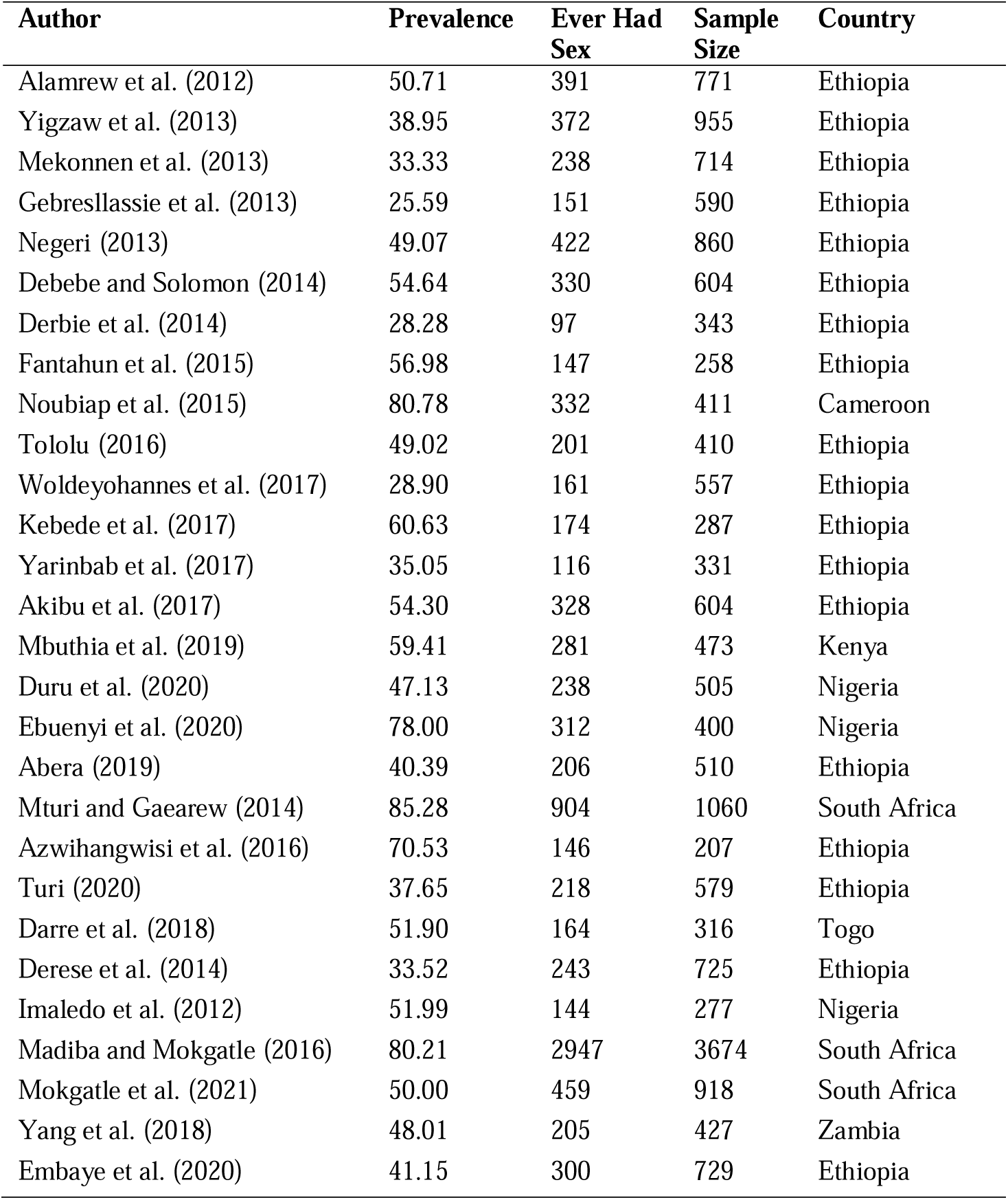
Summary of Studies Included in the Review and Meta-analysis (n = 28)

### Pooled Estimates for Prevalence of Sexual Activity in Sub-Saharan Africa

According to different works of literature in Sub-Saharan Africa, the prevalence of sexual activity in Colleges and Universities ranged from 26.0% to 81%. The estimated pooled prevalence of sexual activity among college and university students was 51.0% (95% CI: 43.0% - 59.0%) (**Figure 2**).

**Figure 1.**
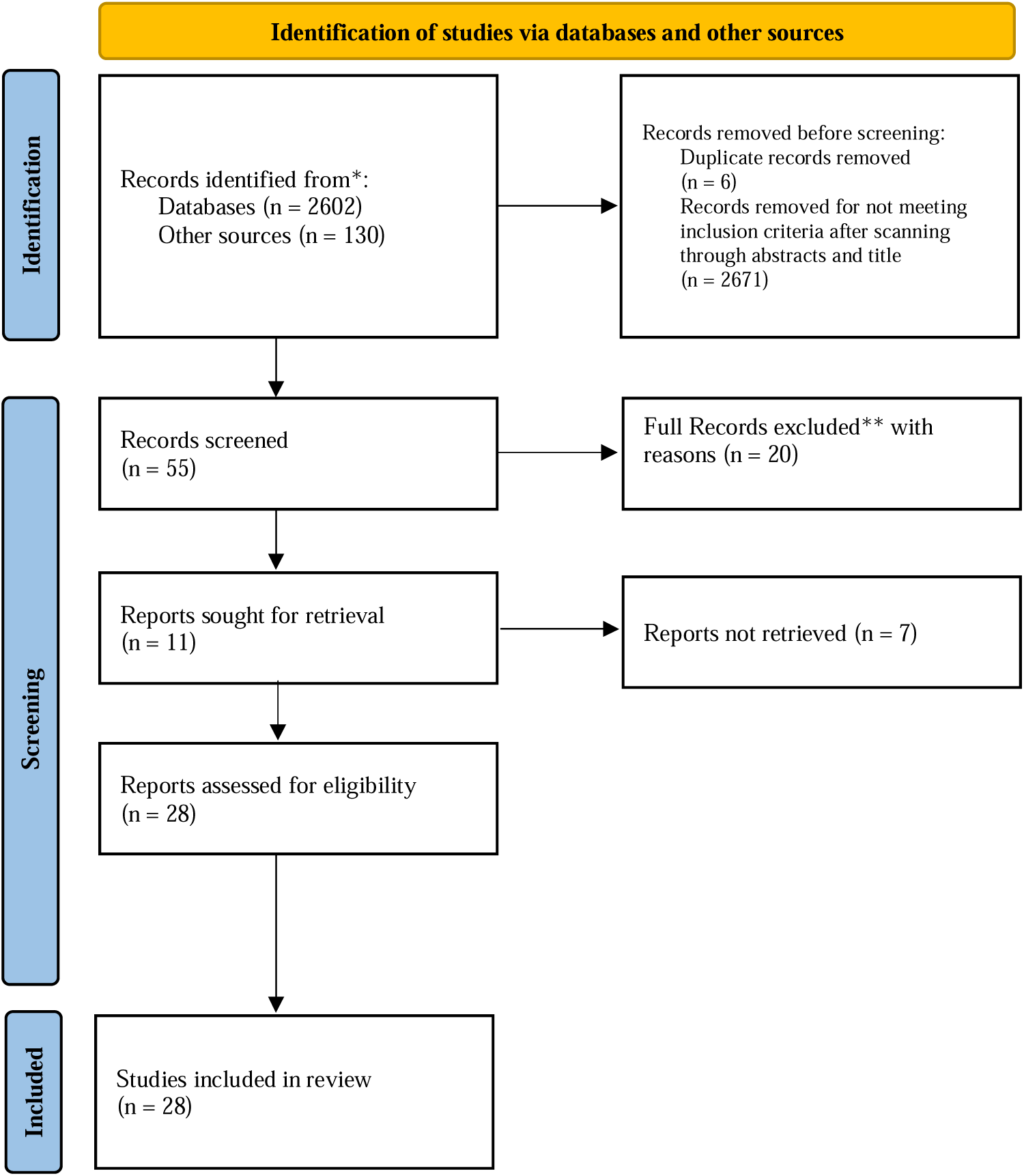
PRISMA Flowchart for the Literature Search

**Figure 2.**
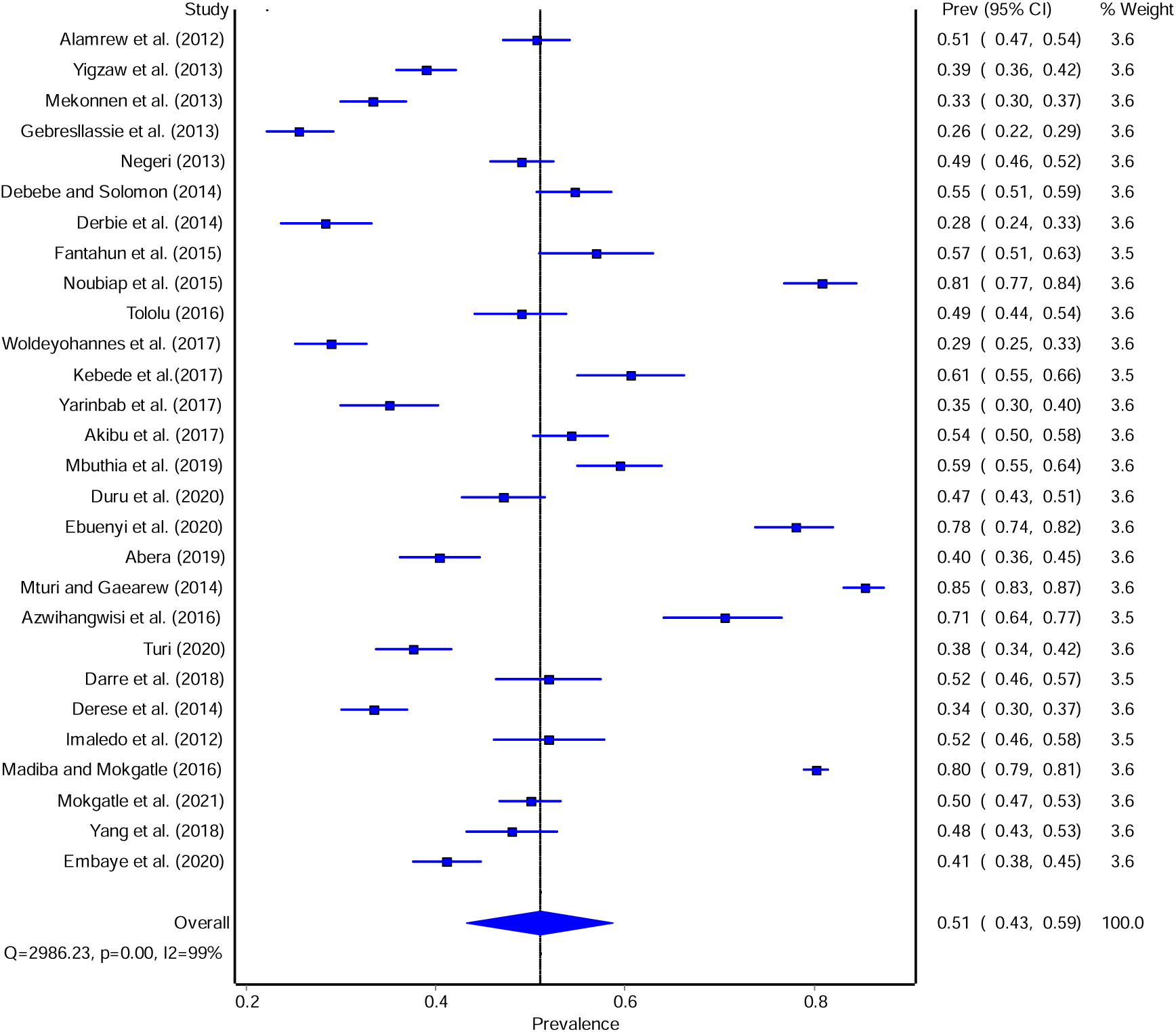
Forest plot for the results of the random-effects meta-analysis of 28 studies on the prevalence of sexual activity among college and university students in Sub-Saharan Africa

### Prevalence of Risky Sexual Behaviours among Students in Colleges and Universities of Sub-Saharan Africa

#### Prevalence of Multiple Sexual Partners

Fifteen (n = 22) studies reported this risky sexual behaviour in their studies. The prevalence of having more than one sexual partner ranged from 12% to 65%. Through the meta-analysis, the pooled prevalence was 36.0% (95% CI: 30.0% - 42.0%) (**Figure 3**).

**Figure 3.**
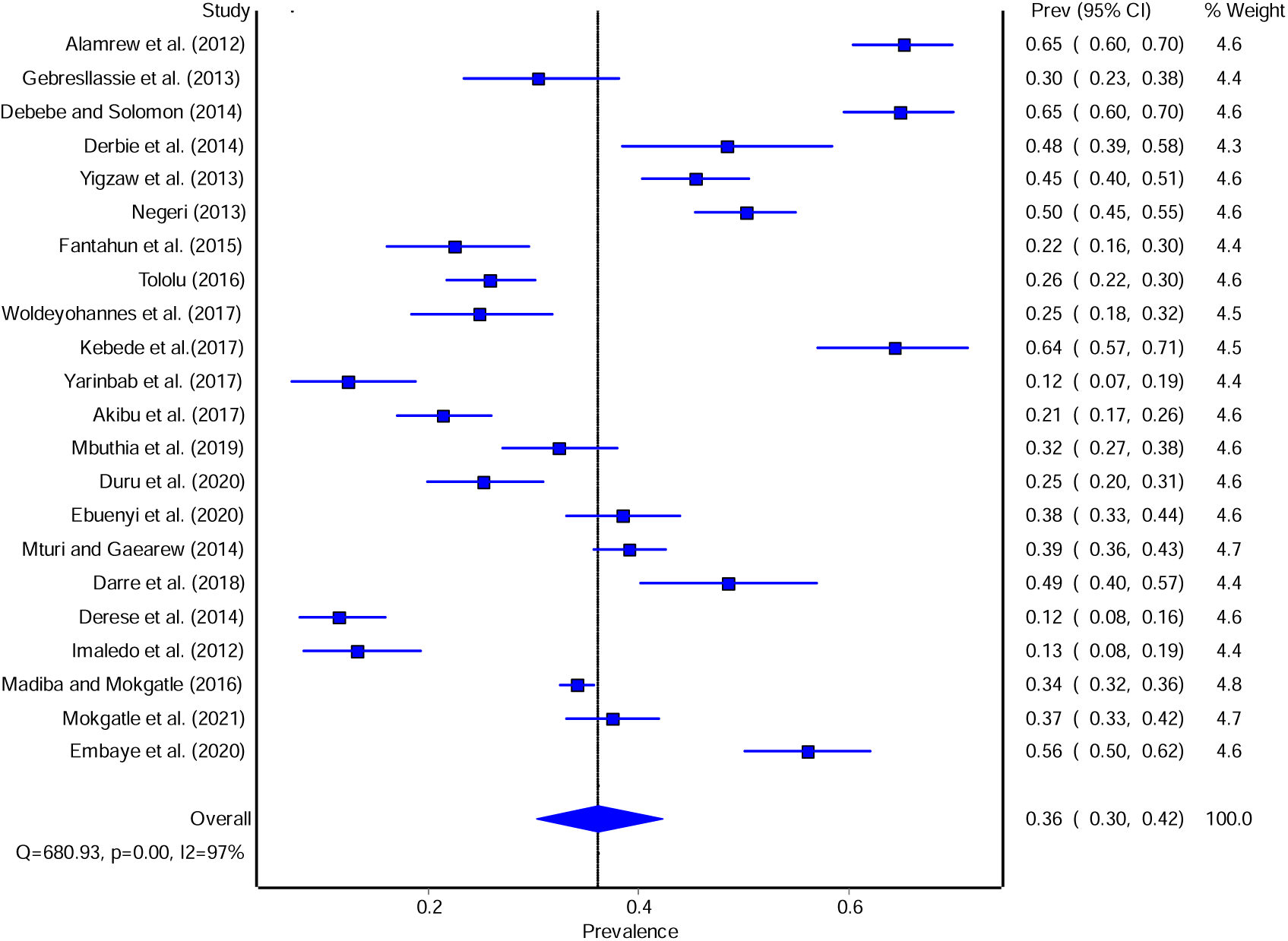
Forest plot for the results of the random-effects meta-analysis of 22 studies on the prevalence of multiple sexual partners among college and university students in sub-Saharan Africa

#### Prevalence of Inconsistent Condom Use

Fourteen studies were combined systematically to produce the pooled prevalence for inconsistent condom use among college and university students in Sub-Saharan Africa. The results revealed that the prevalence ranged from 27.0% to 83.0%. After using the random-effects model, the results obtained showed that the overall prevalence was 53% (95% CI: 46.0% - 61.0%) **(Figure 4)**.

**Figure 4.**
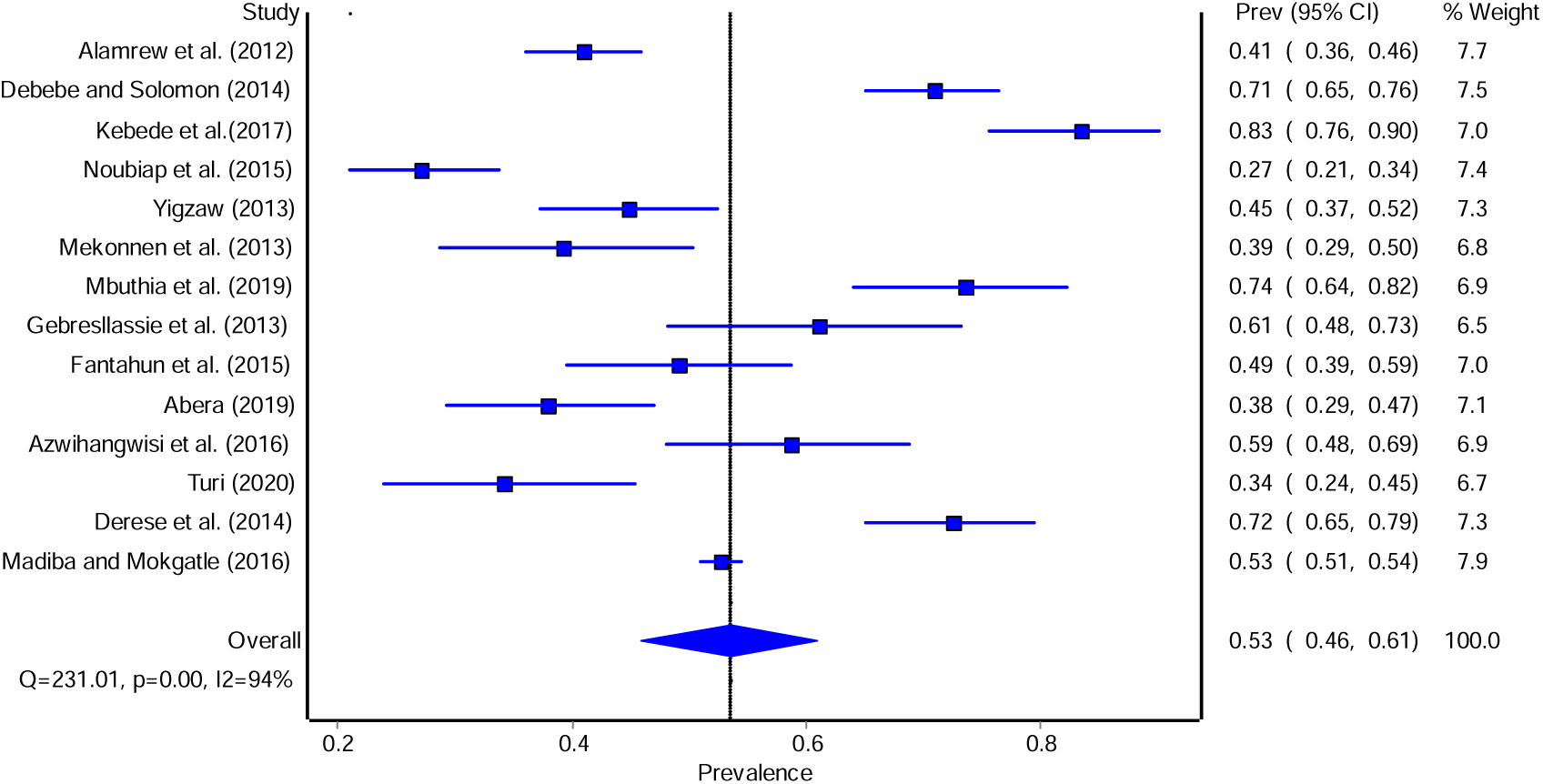
Forest plot for the results of the random-effects meta-analysis of 14 studies on the prevalence of inconsistent condom use among college and university students in sub-Saharan Africa

#### Prevalence of Overall Risky Sexual Behaviors

Seven studies in the reviewed literature reported risky sexual behaviors defined as having at least one of the following:

- Multiple sexual partners
- Unprotected sexual intercourse
- Sexual intercourse under alcohol influence
- Early age at sexual intercourse

From the reviewed literature, this prevalence ranged from 41.0% to 95%. The pooled estimate for the prevalence was found to be 65% (95% CI: 48.0% - 81.0%) **(Figure 5)**.

**Figure 5.**
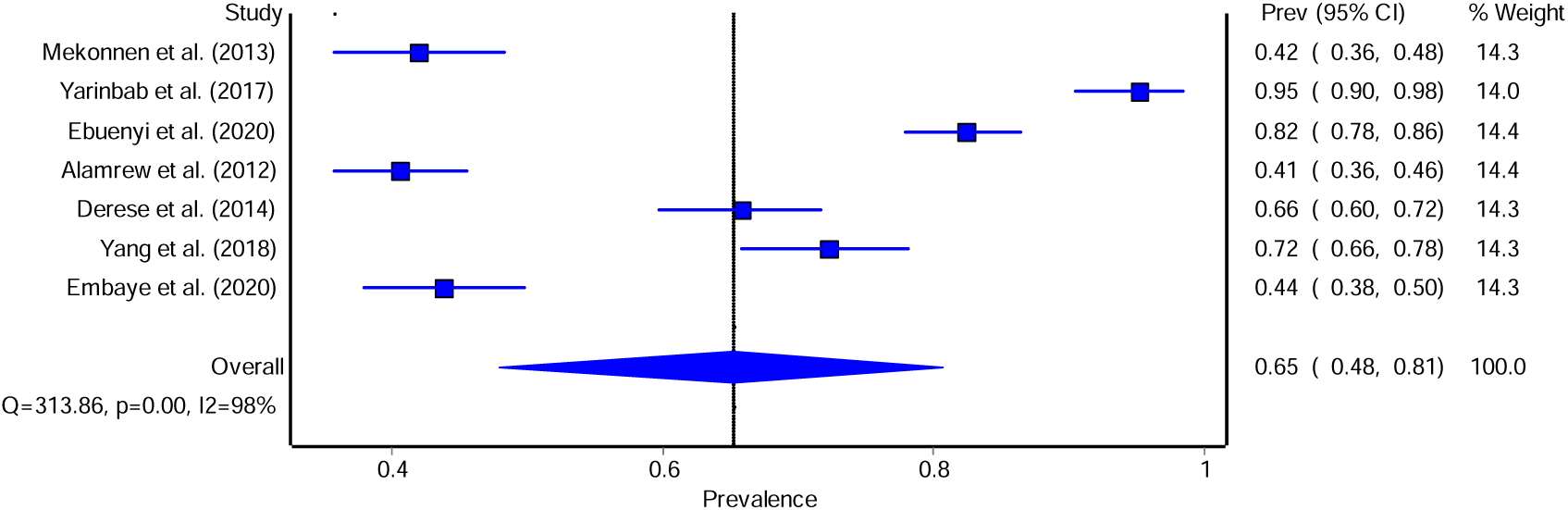
Forest plot for the results of the random-effects meta-analysis of 7 studies on the prevalence of risky sexual behaviour among college and university students in sub-Saharan Africa

### Sex and Consistent Condom Use

Two studies were assessed to determine the effect of sex on consistent condom use. The literature revealed that similar results. Males were more likely to demonstrate consistent condom use compared to female students. This review reveals a non-significant pooled odds ratio of 1.42 (0.70 – 2.88) (**Figure 7**).

**Figure 6.**
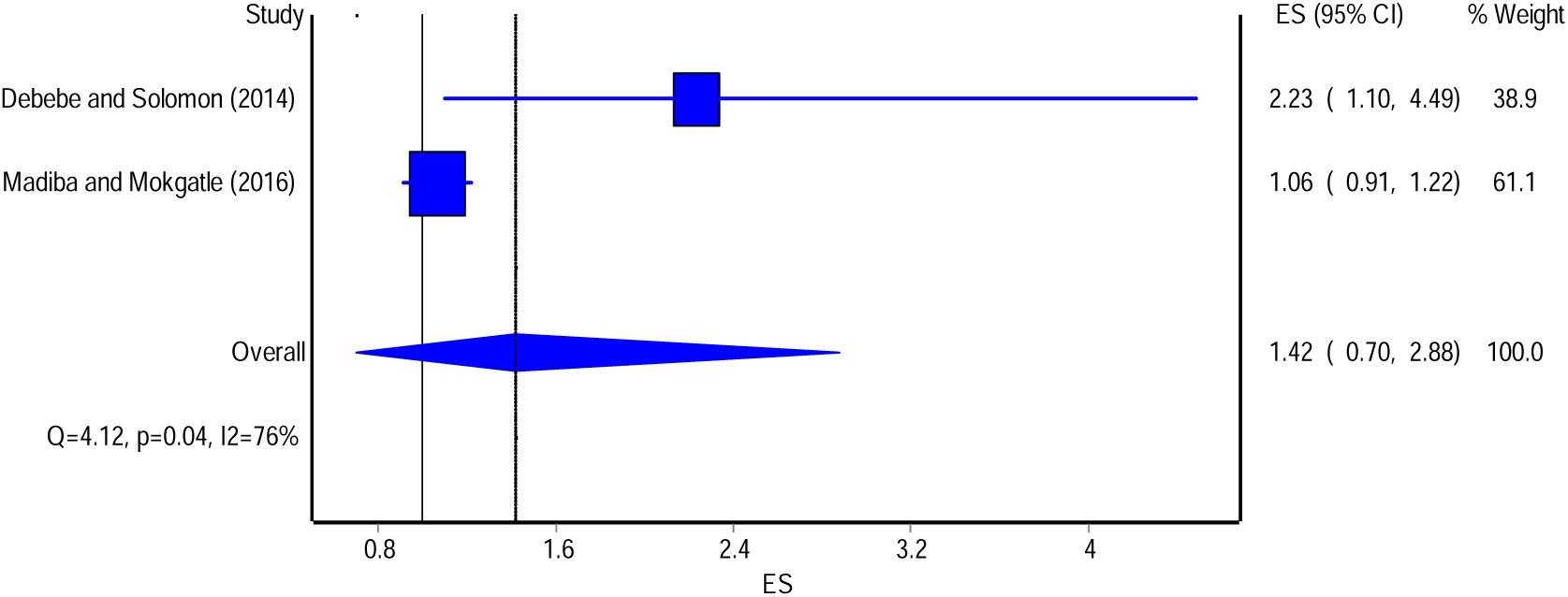
Forest plot for the results of the random-effects meta-analysis of 2 studies on odds ratio for the effect of sex on consistent condom use among college and university students in sub-Saharan Africa

**Figure 7.**
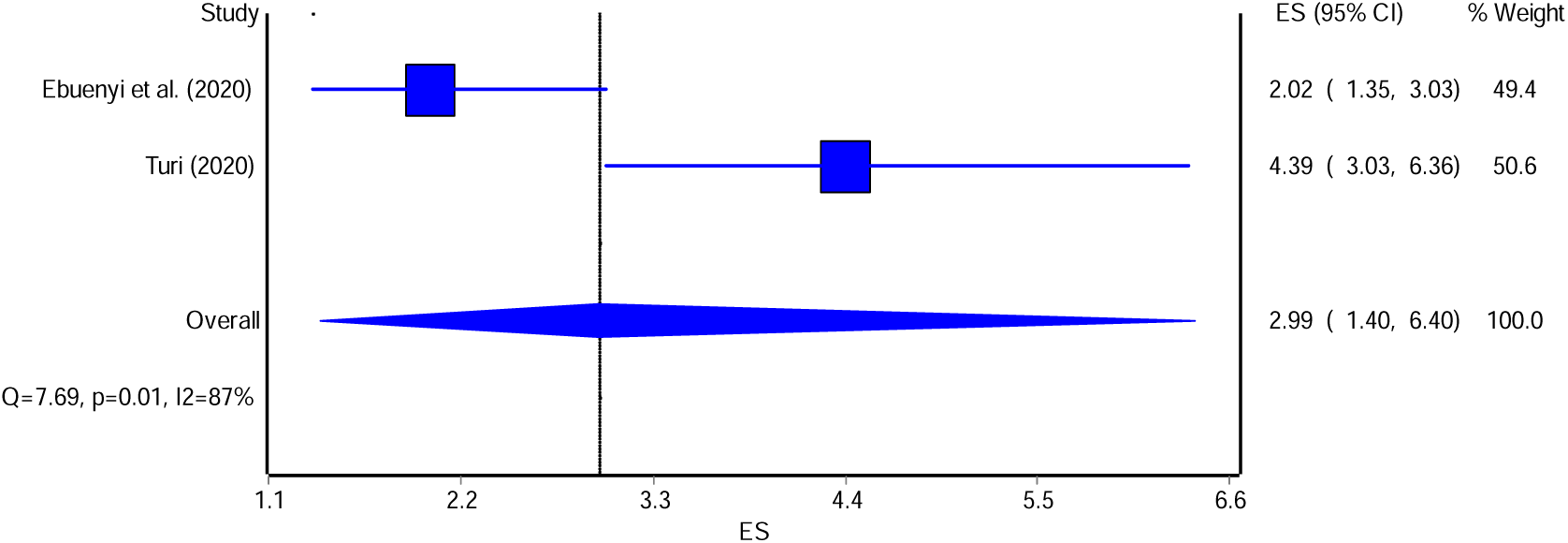
Forest plot for the results of the random-effects meta-analysis of 2 studies on odds ratio for the effect of sex on risky sexual behavior among college and university students in sub-Saharan Africa

### Sex and Risky Sexual Behaviour

This analysis only included two studies. The results indicated that males were 2.99 times more likely to engage in risky sexual behavior than females (OR: 2.99; 95% CI: 1.40 – 6.40) (**Figure 8**).

## DISCUSSION

### Pooled Prevalence Estimate for Sexual Activity and Risky Sexual Behavior

This study found that the estimated pooled prevalence of sexual activity and multiple sexual partners among college and university students was moderately high. Furthermore, more than half of college and university students practiced at least one risky sexual behavior and did not use condoms consistently. This indicates that university and college students continue to practice risky sexual behavior. Thus, urgent interventions must be put in place to discourage risky sexual behavior among university and college students.

These findings are comparable but significantly higher than those reported in a systematic review and meta-analysis conducted in Ethiopia, which indicates a sexual activity prevalence of approximately 41.62 per cent. This difference can be explained by the different inclusion criteria for the studies utilized. This review focused on results across countries part of Sub-Saharan Africa.

This overall observation in developing countries can be explained by the fact that adolescents have become increasingly prone to engage in risky sexual behavior in many developing countries, particularly Sub-Saharan Africa.

The prevalence estimates for sexual activity, multiple sexual partners, unprotected sexual intercourse and sexual activity under alcohol influence among university and college students are a public health concern. The identified risky sexual behavior in this meta-analysis indicate that young people continue to face an increased risk of acquiring STIs and HIV due to an early age of sexual debut, inconsistent condom use, multiple sexual partners and sexual intercourse under the influence of alcohol [2,13]. Failure to use protection against STIs or pregnancy, and choosing poor or risky sexual partners, may continue to directly lead to negative consequences such as STIs, unplanned pregnancy, or sexual assault.

## CONCLUSIONS

Risky sexual behavior remains high in Universities and Colleges in Sub-Saharan Africa. University students continue to engage in risky sexual behavior such as inconsistent condom use and having more than one sexual partner. There is a need to formulate strategies targeting this group, particularly those who drink alcohol and male students, as they demonstrated a much higher risk of engaging in risky sexual behavior. Educational institutions such as colleges and universities in Sub-Saharan Africa must provide and implement adequate health education and behavior change programs targeting men and those who abuse alcohol to reduce risky sexual behavior and the consequences of these practices. Additionally, we would recommend that more studies be done to determine the effect on pornography viewing and risky sexual behavior and the influence on parental sexual education on risky sexual behavior among university and college students.

## Data Availability

All data produced in the present study are available upon reasonable request to the authors

## Ethical considerations

The study did not require access to human subjects as it is a desk-based project. However, all primary research articles included in this study were thoroughly screened to determine if they met the required ethical requirements and principles.

## AUTHOR CONTRIBUTIONS

### AL

- Work conception. Data acquisition, analysis and interpretation.
- Manuscript drafting.
- Approval of final manuscript.
- Accountable for all aspects of the work regarding its accuracy or integrity.

### CC

- Work conception, Data analysis and interpretation.
- Manuscript drafting.
- Approval of final manuscript.
- Accountable for all aspects of the work regarding its accuracy or integrity.

### MZ

- Manuscript drafting.
- Critical revisions for intellectual content.
- Data interpretation
- Approval of final manuscript.
- Accountable for all aspects of the work regarding its accuracy or integrity.

### PJC

- Manuscript drafting.
- Critical revisions for intellectual content.
- Data interpretation
- Approval of final manuscript.
- Accountable for all aspects of the work regarding its accuracy or integrity.

## Acknowledgement

This article is a part of the master thesis submitted to the University of Lusaka in partial fulfilment of the requirements for the degree of Master of Public Health.

